# Lexical Stability of Psychiatric Clinical Notes from Electronic Health Records over a Decade

**DOI:** 10.1101/2022.09.05.22279610

**Authors:** Lasse Hansen, Kenneth Enevoldsen, Martin Bernstorff, Erik Perfalk, Andreas A. Danielsen, Kristoffer L. Nielbo, Søren D. Østergaard

**Author notes:** **Corresponding Author** Lasse Hansen, MSc, Department of Clinical Medicine, Aarhus University, Palle Juul-Jensens Boulevard 175, 8200 Aarhus C, Denmark.

## Abstract

Natural Language Processing methods hold promise for improving clinical prediction by utilising information otherwise hidden in the clinical notes of electronic health records. However, clinical practice—as well as the systems and databases in which clinical notes are recorded and stored—change over time. As a consequence, the content of clinical notes may also change over time, which could degrade the performance of prediction models. Despite its importance, the stability of clinical notes over time has rarely been tested. Therefore, in this study, we examined the lexical stability of clinical notes from the Psychiatric Services of the Central Denmark Region in the period from January 1, 2011, to November 22, 2021 (a total of 14,811,551 clinical notes describing 129,570 patients) by quantifying sentence length, readability, syntactic complexity and clinical content - and estimating changepoints in these metrics. We find lexical and syntactic stability over time, which bodes well for the use of Natural Language Processing for predictive modelling in clinical practice.

## Introduction

In medical care, electronic health record (EHR) systems are used to document all patient-related information. The type of recorded information is highly heterogeneous, and covers lab results, diagnoses, administered medication, as well as subjective (reported by the patient) and objective (findings made by the healthcare professionals) descriptions of behaviour and symptoms in the form of text in clinical notes^1^. Notably, the clinical notes are not only a site for collection—but also of aggregation and synthesis—of clinical information^2^. Especially in psychiatry, important clinical information is often documented as text^3^.

As the volume of information increases in EHR systems, it may become increasingly harder for health care professionals to make meaningful decisions based on this information^4^. In these situations, decision-support based on analysis of the EHR data can guide clinicians towards relevant information and suggest courses of action^5^. Whereas structured numerical data can relatively easily be aggregated with conventional statistical descriptors (e.g. minimum and maximum values, means, medians etc.), extracting relevant information from free-form clinical text requires a different approach^6^. In recent years, however, the field of natural language processing (NLP) has made marked progress^7–9^, and textual information is no longer outside the purview of, e.g., machine learning algorithms. Indeed, the utility of written text for prediction models in healthcare has been investigated in multiple studies and has been found to improve performance^10–15^. However, the stability of this predictive performance relies on the EHR source’s stability, including the text in the clinical notes. Specifically, the performance of prediction models tends to degrade markedly the more the data used for prediction differs from that used for training^16^. Formally, in the case of distribution shift or drift, the assumption of identically distributed data no longer holds. This can lead to spurious or highly time-dependent patterns in prediction, which are unlikely to generalise^17^.

Hospital services using EHR are highly dynamic; for instance, clinical procedures, treatments, diagnostic criteria, the distribution of tasks between healthcare professions, and EHR systems tend to change substantially over time. The EHR information contained in written clinical notes may be particularly prone to changes over time, but—to our knowledge—the stability of the written clinical notes from EHR is rarely tested. Therefore, we aimed to assess the lexical stability over time of written clinical notes from a large psychiatric hospital system. More specifically we aimed to answer the question of lexical stability using data from the Psychiatric Clinical Outcome Prediction (PSYCOP) cohort^18^, which contains EHR clinical notes covering almost 130,000 patients and more than 14,000,000 clinical notes over the ten years from 2011 to 2021. We employed change point detection models to examine whether i), the use of words describing psychopathology (symptoms) in clinical notes in the EHR is stable over time and ii) if sentence length, readability, syntactic complexity, and usage of different types of clinical notes is stable over time. The methods employed and the results obtained may serve as a benchmark for future studies testing the lexical stability of clinical notes from other hospital systems.

## Results

Tables 1 and 2 show dataset characteristics related to the number of patients and note types. The dataset contains data from a total of 129,570 patients, of whom 66,575 (51%) are female, and is representative of all age groups. The dataset contains more than 1 billion tokens and 14.8 million notes across the 17 selected note types, with a mean note length of 71.5 tokens.

**Table 1:** Demographic characteristics of the cohort. The table shows the number of unique patients grouped by their last ICD-10 diagnosis, age group at the first visit, and sex. * if less than ten patients in a group, the cell count is set at <10 to avoid the identification of individuals.

| Age | <18 |  | >18 |  |
| --- | --- | --- | --- | --- |
| Last ICD-10 diagnosis | Females | Males | Females | Males |
| F00-F09 Organic mental disorders | <10* | <10* | 5235 | 3696 |
| F10-F19 Mental and behavioural disorders due to psychoactive substance use | 40 | 113 | 967 | 2501 |
| F20-F29 Schizophrenia, schizotypal and delusional disorders | 361 | 245 | 3275 | 4459 |
| F30-F39 Mood [affective] disorders | 850 | 407 | 12355 | 7621 |
| F40-F48 Neurotic, stress-related and somatoform disorders | 3184 | 1654 | 10865 | 8505 |
| F50-F59 Behavioural syndromes associated with physiological disturbances and physical factors | 1158 | 132 | 1910 | 553 |
| F60-F69 Disorders of adult personality and behaviour | 383 | 38 | 3469 | 1129 |
| F70-F79 Mental retardation | 244 | 408 | 790 | 904 |
| F80-F89 Disorders of psychological development | 1711 | 5074 | 427 | 852 |
| F90-F98 Behavioural and emotional disorders with onset usually occurring in childhood and adolescence | 3773 | 7827 | 2713 | 3714 |
| Other diagnoses | 3168 | 3432 | 9693 | 9723 |

**Table 2:** Number and length of notes by note type. Sorted in descending order based on total number of tokens.

| Note type | Number of notes | Total tokens | Mean tokens per note |
| --- | --- | --- | --- |
| Observation of patient, Psychiatry | 2,972,943 | 202,733,055 | 68.19 |
| Current mental state | 2,227,647 | 354,778,113 | 159.26 |
| Medication | 1,837,916 | 70,759,048 | 38.5 |
| Objective mental state | 1,485,799 | 66,716,343 | 44.9 |
| Appointments, Psychiatry | 1,404,610 | 38,665,226 | 27.53 |
| Conversation with treatment aim | 967,541 | 77,400,486 | 80.00 |
| Current social function, Psychiatry | 710,314 | 48,003,618 | 67.58 |
| Current somatic, Psychiatry | 513,777 | 19,739,080 | 38.42 |
| Telephone consultation | 426,126 | 24,772,013 | 58.13 |
| Plan | 424,641 | 21,544,611 | 50.74 |
| Reason for contact | 421,300 | 20,679,254 | 49.08 |
| Patient note | 419,942 | 28,029,332 | 66.75 |
| Conclusion | 398,857 | 35,871,674 | 89.94 |
| Telephone note | 246,163 | 16,875,687 | 68.55 |
| Prescription | 199,870 | 3,852,471 | 19.27 |
| Objective, somatic | 99,915 | 4,746,572 | 47.51 |
| Semi-structured diagnostic interview | 54,190 | 24,425,900 | 450.75 |
| Total | 14,811,551 | 1,059,592,483 | 71.50 |

### Stability across all notes and words describing psychopathology

As seen in Figure 1, the mean number of tokens, mean dependency distance, automated readability index, and novelty on the aggregate level (across all clinical notes or all words describing psychopathology) were relatively stable over the course of the study. No changepoints were found in any of the groups. However, the mean number of tokens increased steadily over time.

**Figure 1:**
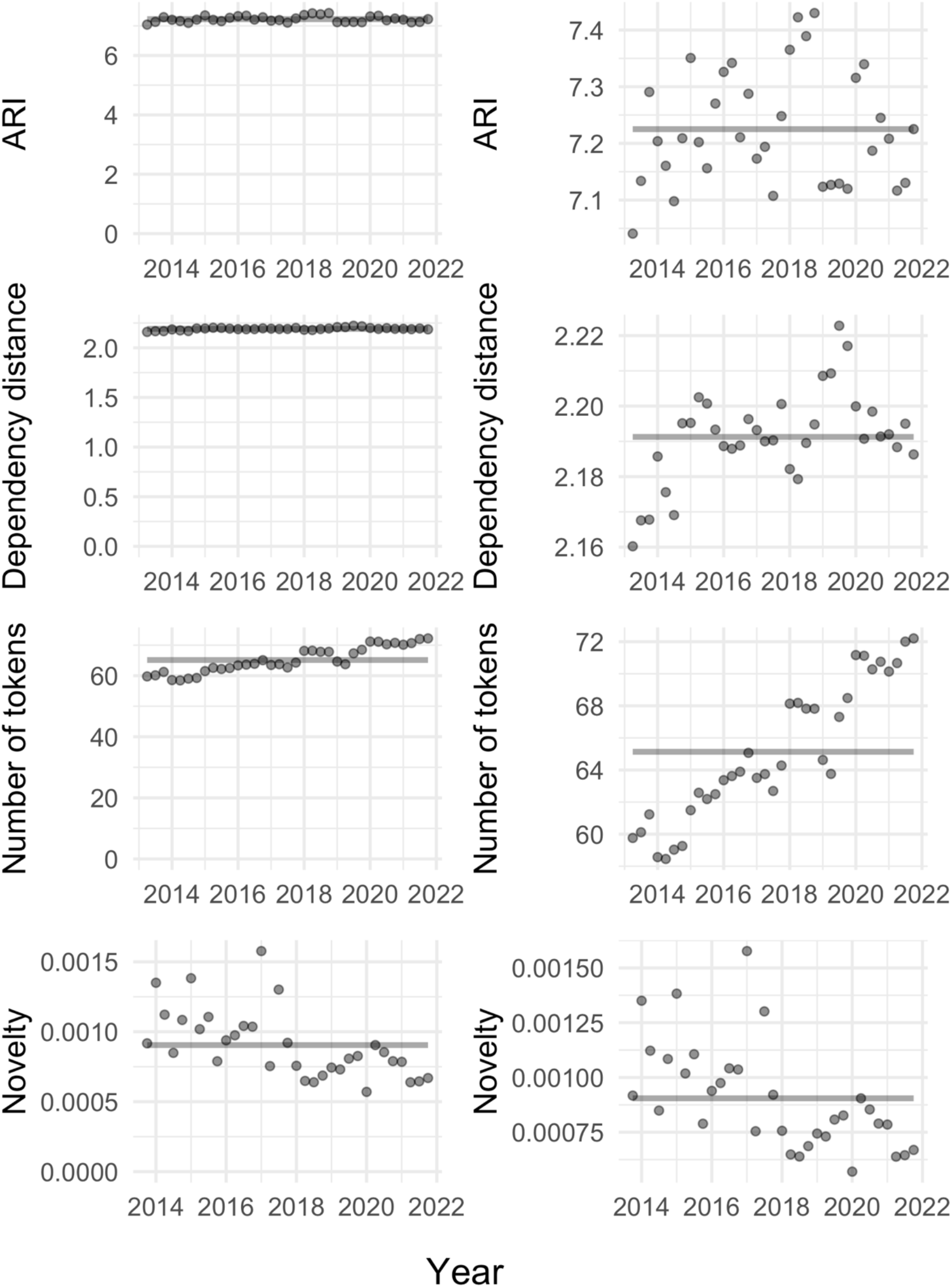
Time series for ARI, mean dependency distance, the mean number of tokens for the aggregated clinical notes (top 3 rows), and novelty across all psychopathological keywords. The grey line indicates the estimated changepoint segments (none found for any groups). The left column shows the data with the y-axis going to 0, and the right column shows the data with the y-axes allowed to vary.

#### Analysis i: Stability in the use of words describing psychopathology

As shown in Figure 2, changes in novelty were found in two out of the 12 (diagnostic) categories of words describing psychopathology (ICD-10 codes F0-F9, objective description, and aggregated words), namely F1 (Substance abuse) and F3 (Mood disorders). The novelty was low across the word categories, which indicates that the distributions are generally stable over time. Visual inspection of the identified changepoints in F1 – Substance abuse (Supplementary Figure 5) suggests that these are artefacts rather than actual changepoints as they do not map onto any visually salient changes. The novelty for words describing psychopathology related to substance abuse was extremely low, which suggests that the distribution is particularly stable. For F3 – Mood disorders, the changepoint reveals a slight rise in novelty in 2020 Q2, followed by a consistent drop (see Supplementary Figure 5). Figure 3 shows that this rise was mainly driven by decreased use of “nihilistic delusions” and “grandiose delusions” as well as increased use of “dysthymia”.

**Figure 2:**
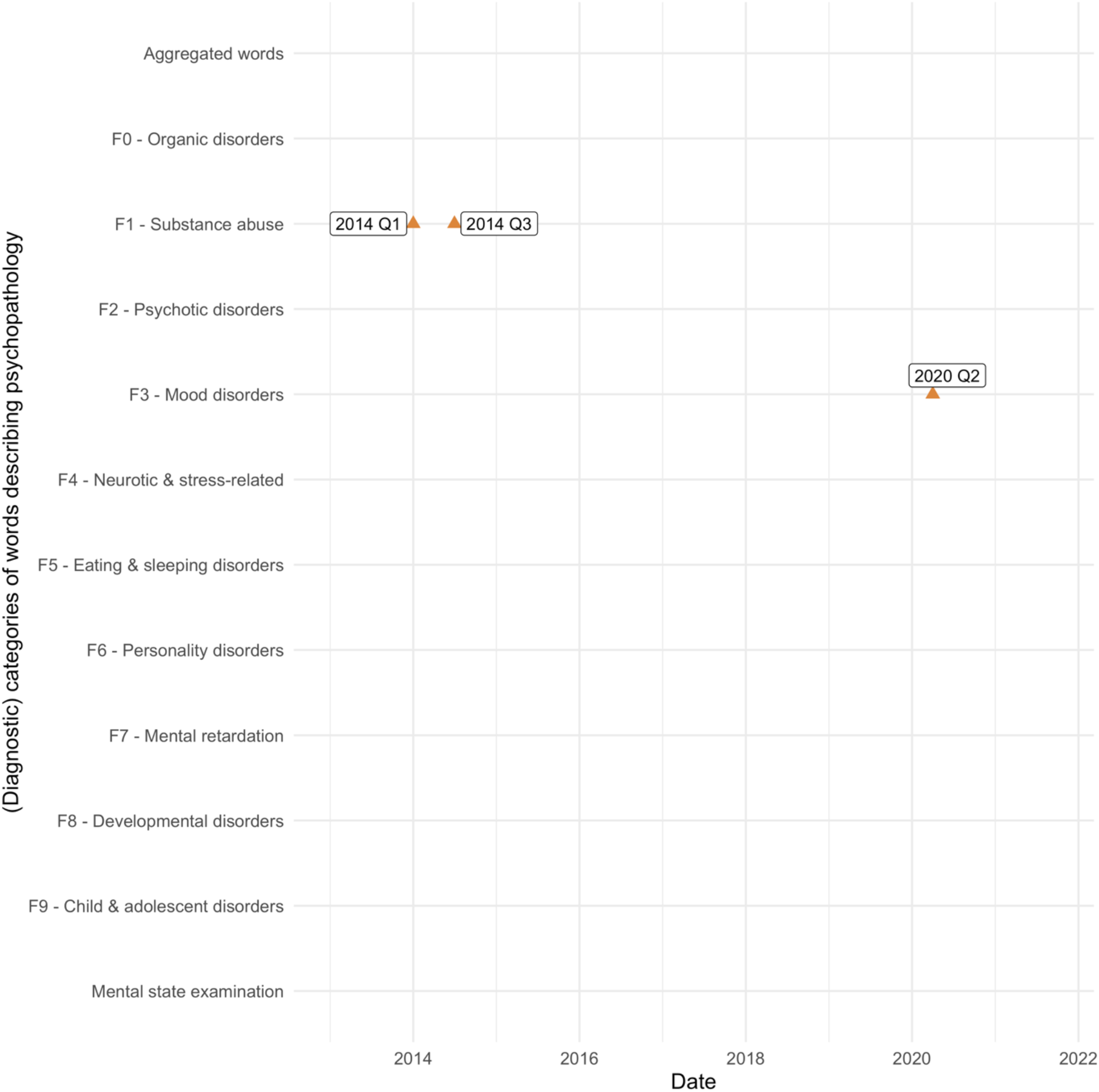
Location of changepoints on novelty for each clinical keyword category.

**Figure 3:**
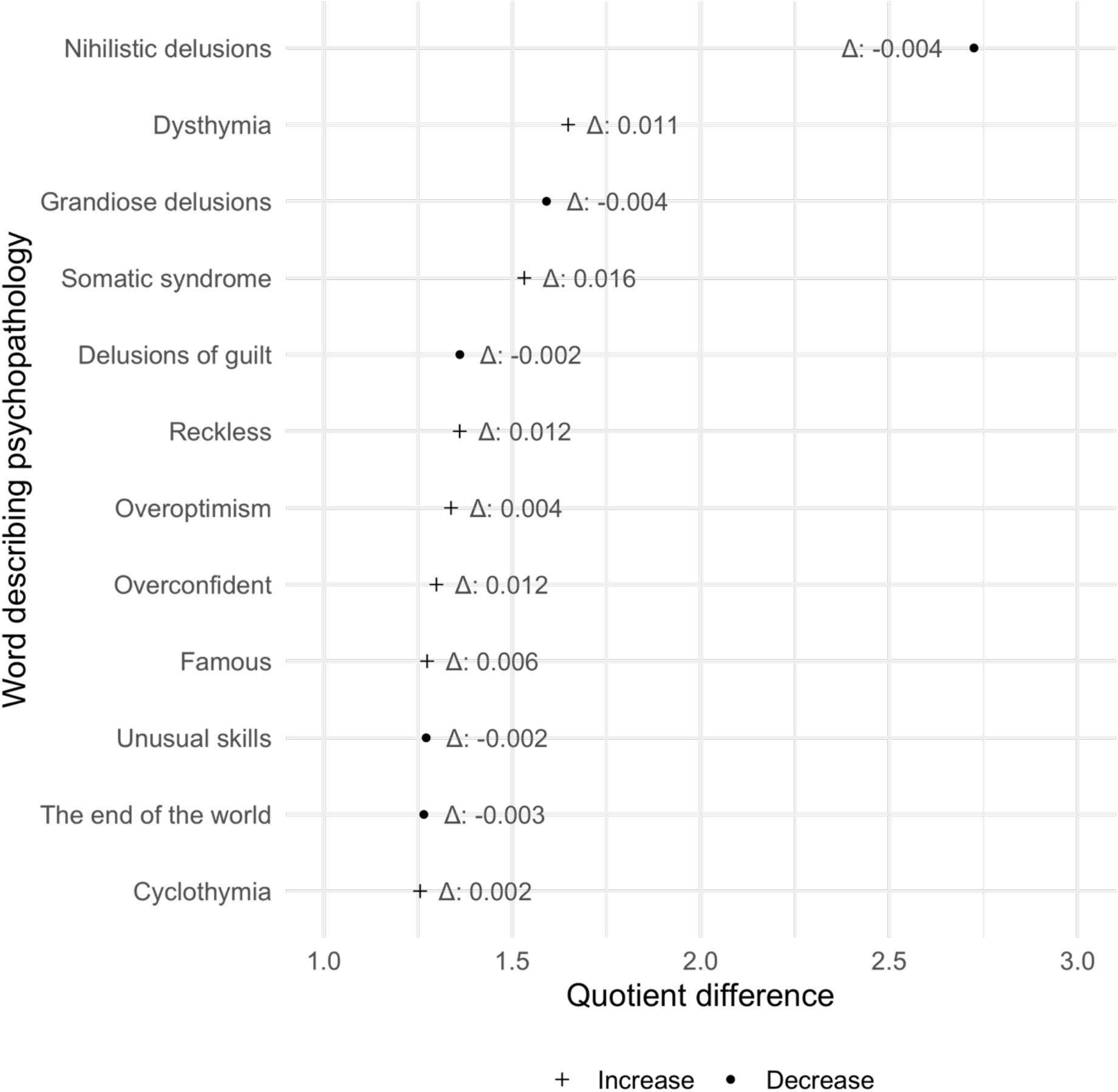
The 12 words describing psychopathology with the largest relative change at the 2020 Q2 changepoint for F3 – Mood disorders. The x-axis shows the quotient difference between the mean of the two previous time points (novelty window) to the time point of interest (1 = no difference). The shape indicates whether there was an increase or decrease in word use. The text next to the label denotes the actual difference in means, i.e., a change of 0.012 means that the use of a word rose to comprise an additional 0.012% of all words in the F3 – Mood disorder category (the 68 words in the F3 – Mood disorder category are available in Supplementary Materials Table 2) in 2020 Q2 compared to the previous two quarters.

#### Analysis ii: Stability of sentence length, readability metrics, and syntactic complexity of clinical notes

Figure 4 shows the changepoints for the mean number of tokens, mean dependency distance, Automated Readability Index (ARI), and proportion of total notes for each note type. Across the 17 note types and four categories of variables, a single changepoint was identified for the number of tokens, two for dependency distance, four for ARI, and seven for note-type proportion (a total of 14 changepoints). The changepoints were concentrated mainly around the year 2020; however, most of the estimated changepoints were due to deviations from a linear trend or represented artefacts caused by reduced usage of the note type due to administrative changes (see the discussion for further explanation). No changepoints were found for eight of the note types, nor when analysing all notes together (aggregate).

**Figure 4:**
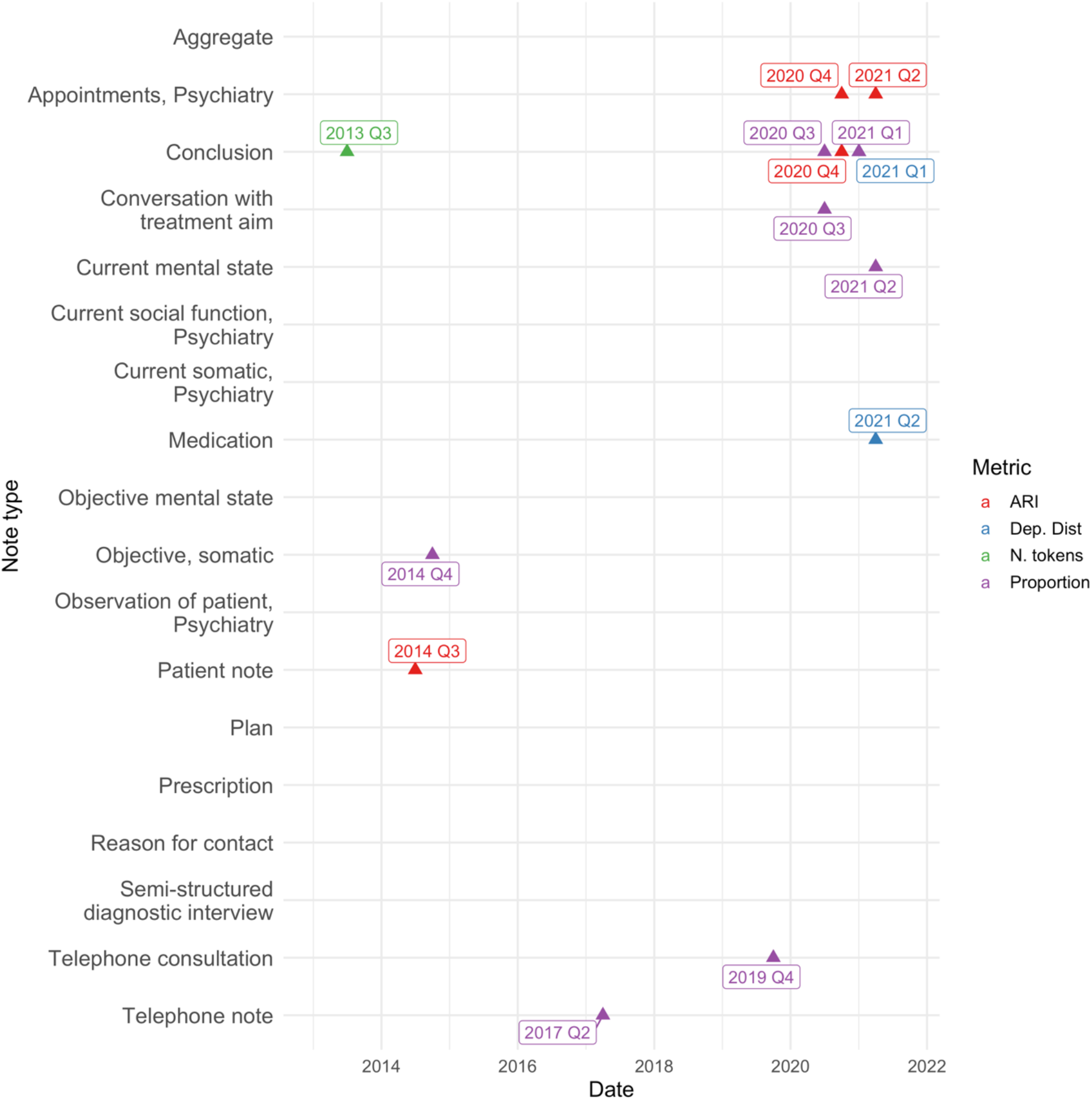
Location of the 14 changepoints in the mean number of tokens, dependency distance, Automated Readability Index (ARI), and proportion of total notes for each note type. The colours indicate which metric the changepoint occurred in.

Supplementary Figures 1-4 show the time series for the mean number of tokens, dependency distance, automated readability index (ARI), and proportion of total notes for all note types with the output from the changepoint model overlaid. Half of the estimated changepoints (7 out of 14) were related to changes in how large a proportion of the total notes the note type made up. ‘Conclusion’ had the most significant change by practically falling out of use after 2020 Q3, going from comprising a mean of 2.7% of notes from 2013 to 2020 Q2 to only comprising a mean of 0.3% afterwards. Therefore, the changes in ARI and dependency distance in 2020 Q4 and 2021 Q1, respectively, are likely to be artefacts driven by a very low number of examples (see the discussion for further explanation). The proportion of notes stemming from “Phone consultations” rose from comprising 2.5% of notes on average in 2014-2019 Q2 (pre COVID-19 pandemic) to 3.9% afterwards (during the COVID-19 pandemic). The proportion of “Telephone notes” also rose during the COVID-19 pandemic. “Conversation with treatment aim” saw a drop in usage in 2019, from comprising approximately 6.5% of notes to 5%. The change in “Current mental state” seems to reflect a minor deviation from a linearly decreasing trend after 2018. “Objective, somatic” saw a halving in usage from comprising a mean of 1.2% of notes before 2014 Q4 to a mean of 0.5% afterwards.

The changepoints in the ARI (4 out of 14) include a slight increase in trend for ARI for “Appointments, Psychiatry” in 2020, and a slight outlier in 2014 Q2 for “Patient note”, with a mean ARI of 9.5 compared to a mean of 8.5 in the previous year. The changepoint in ARI for “Conclusion” in 2020 Q3 is due to reduced usage of the “Conclusion” note type. All dependency distance-related changepoints (2 out of 14) seem to be related to reductions in the use of the note type. As with ARI, the 2020 Q4 “Conclusion” changepoint is an artefact of reduced usage. The 2021 Q1 changepoint in “Medicine” appears to reflect a degree of variability over time rather than a salient changepoint. Only a single changepoint (1 out of 14) was identified in the mean number of tokens per note in the “Conclusion” note type in 2013 Q2. Visual inspection of Supplementary Figure 1 revealed the changepoint to be a minor deviation from a linearly increasing trend.

## Discussion

This study investigated the lexical stability of clinical notes in EHR from a large cohort of patients attending psychiatric services in Denmark. Analyses of changepoints in i) the use of words describing psychopathology – as well as in ii) note length, readability, syntactic complexity, and usage of the clinical notes found the EHR content to generally be stable over time. Notably, no changepoints were found on the aggregate level, that is, when analysing all notes and all words describing psychopathology together. Out of 2988 data points, 17 (0.6%) possible changepoints were identified. We found most changepoints to be related to the discontinuation of a specific note type (“Conclusion”) or from increases in the use of virtual communication during the COVID-19 pandemic. Changepoints for words describing psychopathology were only found in the ICD-10 F3 – Mood disorders category and were likely caused by a change in the patient presentation during COVID-19. A slow distribution shift was, however, observed in the note length of some specific note types (“Current mental state”, “Current social, Psychiatry”, “Current somatic, Psychiatry”, “Patient note”, “Medication”, “Plan”, “Semi-structured diagnostic interview”), which was also apparent on the aggregate level. These changes were not detected by changepoint analyses yet might gradually degrade the performance of prediction models over time. This underlines the importance of thoroughly evaluating the performance of clinical prediction models over time and continuously monitoring them after deployment to ensure model quality.

The most prominent change was found in the use of the “Conclusion” note type, which dropped from comprising a mean of 2.7% of notes before 2020 Q3 to only comprise a mean of 0.3% afterwards. Conversations with officials from the Business Intelligence Office of the Central Denmark Region, which administers the EHR, revealed the large drop in usage of “Conclusion” to be caused by the introduction of a new note type in December 2020, namely “Evaluation/conclusion”, which was intended to replace “Conclusion”. This change was not otherwise documented and implies that one should be cautious if using “Conclusion” as a point of analysis and highlights the importance of evaluating the stability of EHR data records before use for research purposes.

Furthermore, the analysis revealed a rise in the proportion of “Phone consultations” and “Telephone notes” during the COVID-19 pandemic, which is compatible with the regional policy on replacing physical meetings with patients with telephone- or video calls to avoid the spread of the coronavirus. In fact, the build-up of changepoints around the year 2020 is likely influenced by the COVID-19 pandemic. Previous research from the Central Denmark Region has identified changes to language use in the clinical notes during the pandemic^19^, exacerbation of psychopathology amongst patients^20,21^, and large fluctuations in the number of referrals to the psychiatric services^22^, which has likely led to a different patient mix during the pandemic. These factors are likely contributors to the relative instability from 2020-2021.

The analysis of the use of words describing psychopathology suggests that there are changes in the use of terms related to mood disorders in 2020 Q2, after which novelty immediately dropped. Due to the timing of this changepoint, it may be caused by the COVID-19 pandemic – as a consequence of either the logistic changes described in the paragraph above (e.g. physical meetings replaced by phone- or video consultation) or of changes in psychopathology due to the COVID-19 pandemic^20,22^. However, as described in the Methods section, the calculation of novelty is sensitive to relative changes in the use of words describing psychopathology rather than absolute changes. Therefore, due to the rarity of the words driving the change in novelty for F3 – Mood disorders, a small increase in the number of patients with these symptoms can lead to relatively large changes in novelty. These changes in novelty are likely of minor importance, supported by the fact that the overall novelty in the present dataset is low compared to other sources, such as literature or Danish news^23,24^. For instance, for the group of words describing psychotic- and mood disorders, the mean novelty was around 0.001, whereas the novelty of literature or Danish news rarely drops below 0.2. Although different representations were used to calculate novelty (topic model for news and word frequencies in the present case), we argue that the novelty in the use of words describing psychopathology is particularly low.

### Limitations

There are several limitations to this study. First, using frequencies of words describing psychopathology as the basis for novelty calculation will not identify the emergence of new terms or find shifts in writing style. Novelty can be calculated on any distribution and is often based on topic modelling (e.g. Latent Dirichlet Allocation (LDA)^25^), which might provide a higher-level representation of the data. Because LDA and other latent variable models are known to perform badly on short texts^26^, we chose to use the relative words usage describing psychopathology over time to calculate novelty, as this is invariant to the length of the clinical notes. Another option for short texts is to use contemporary neural embedding techniques such as Transformer-based topic modelling^27^, but these are not feasible to use without GPU access, which was not available to us on the server of the Central Denmark Region at the time of the study. Second, novelty is based on relative changes, not absolute changes. This means that our analysis is sensitive to changes in rare psychopathological keywords as they are likely to fluctuate more than commonly used words. This might increase the risk of finding artefacts due to changes in the patient population. Third, to identify changes in writing style and writing conventions, one could use methods for out-of-distribution detection such as perplexity of the texts over time^28^. This, however, requires the use of a GPU as well as a generative language model, neither of which were available to us. Fourth, changepoint detection models are not sensitive to slow distribution shifts and drifts but rather to changes in means or variance that persist over some time. As a consequence, our method will identify major breaks, such as the discontinuation of the “Conclusion” note type but will not find distribution shifts, such as the increase in note length, without manual analysis and inspection. Fourth, there are several different formulas for calculating readability. We chose to use ARI as it is simple to implement, coupled with the fact that it had the highest mean correlation (0.93) to the other readability metrics available in the *textdescriptives* python package^29^. Given the high correlation, it is reasonable to expect other readability metrics to produce very similar results.

### Conclusion

In a large body of psychiatric clinical notes, the mean length of notes, readability, syntactic complexity, note type distribution and usage of words describing psychopathology were generally stable. The discontinuation of the “Conclusion” note type was the cause of most of the changepoints detected. There were changes in novelty in some groups of words describing psychopathology, but the overall novelty was relatively low, suggesting consistency in the content of the clinical notes over time. Taken together, these findings suggest that prediction models can be trained on the content of clinical notes without fitting to idiosyncrasies of a specific time period. However, one should be cautious of gradual changes in the data distribution, such as an increase in the average note length.

## Methods

### Data source

We used data from an updated version of the Psychiatric Clinical Outcome Prediction (PSYCOP) cohort^18^. Specifically, we analysed all EHR clinical notes from all patients with at least one contact with the Psychiatric Services of the Central Denmark Region in the period from January 1, 2011, to November 22, 2021, covering 129,570 patients and 14,811,551 notes. In the EHR, the clinical notes are labelled according to their content, e.g., “current mental state”, “current social state”, or “medication” (all labels are listed in the Supplementary Information). For the present study, we selected 17 note types, which are among the most widely used and rich in text. See Supplementary Table 1 for a description of the note types.

### Data Processing

For the analysis (i) of the stability of descriptions of psychopathological symptoms over time, we derived a set of 365 words describing psychopathology from the Present State Examination^30^. The 365 words describing psychopathology were grouped by the International Statistical Classification of Diseases and Related Health Problems 10^th^ Revision (ICD-10) diagnostic category which the symptom best described^31^. Subsequently, we counted how many times (summed at the quarterly level) each of the words describing psychopathology—grouped by ICD-10 diagnostic category as well as a category for words used in a mental state examination—occurred in each of the 17 types of clinical notes. Words that occurred less than ten times in five or more quarters were removed to reduce the sparsity of the keyword matrix and thereby increase the robustness of the analysis. A total of 56 keywords were removed due to this criterion (see Supplementary Table 2 for a full list of included and excluded keywords). For each diagnostic group (ICD-10: F0 to F9 and mental status examination), as well as for all words describing psychopathology together (aggregated words describing psychopathology), the counts of words describing psychopathology were transformed to proportions to calculate the *novelty* of each quarter to the previous two quarters. *Novelty* is a measure of windowed relative entropy that expresses information novelty as a difference in content from the past and is further described (under “Metrics”) below.

For the analysis (ii) of the stability of sentence length, readability metrics, syntactic complexity, and usage of the clinical notes over time, we extracted the mean note length, the mean Automated Readability Index (ARI)^32^, and mean dependency distance from each note type using the *spaCy* v3.1.1^33^ and *textdescriptives* v1.0.6^29^ Python libraries and the *spaCy da_core_news_lg* model. For these analyses, we randomly sampled 10% of the notes to reduce computational costs. The proportion of total notes was calculated by dividing the number of notes of each type by the total number of notes (out of the 17 note types). Each metric was aggregated as the quarterly mean, and the resulting time series of quarterly means were then analysed using changepoint detection algorithms. The aggregation was carried out to reduce noise and computational costs. The metrics, as well as the changepoint detection analysis, are described in more detail below.

Data from before 2013 was found to be highly noisy and therefore excluded from the main analyses. The noise was largely due to the gradual implementation of a new EHR system in the region starting in 2011^18,34^. See Supplementary Figure 7 for more details.

### Metrics

#### Novelty

Novelty measures the average divergence between the present keyword distribution *s*^*(j)*^ and past keyword distributions *s*^(*j*−1)^, *s*^(*j*−2)^, *s*^(*j*−*w*)^ in window *w:*

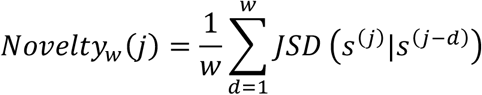

Where JSD is the Jensen-Shannon Divergence:

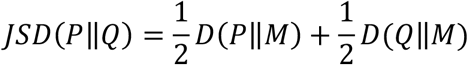

Where *P* and *Q* are probability distributions, 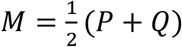 and *D* is the Kullback-Leibler Divergence.

Novelty can be interpreted as the degree of relative entropy or ‘surprise’ in the distribution based on the recent past. The novelty will rise if word usage changes, but if the change becomes more permanent, the novelty will fall again as it becomes the new normal. Many continual changes, i.e., low predictability, cause high novelty, and conversely, few changes cause novelty to be low. A visual representation of how changes affect novelty is shown in Figure 5. Code for calculating novelty can be found on the following Github repository: https://github.com/Aarhus-Psychiatry-Research/lexical-stability

**Figure 5:**
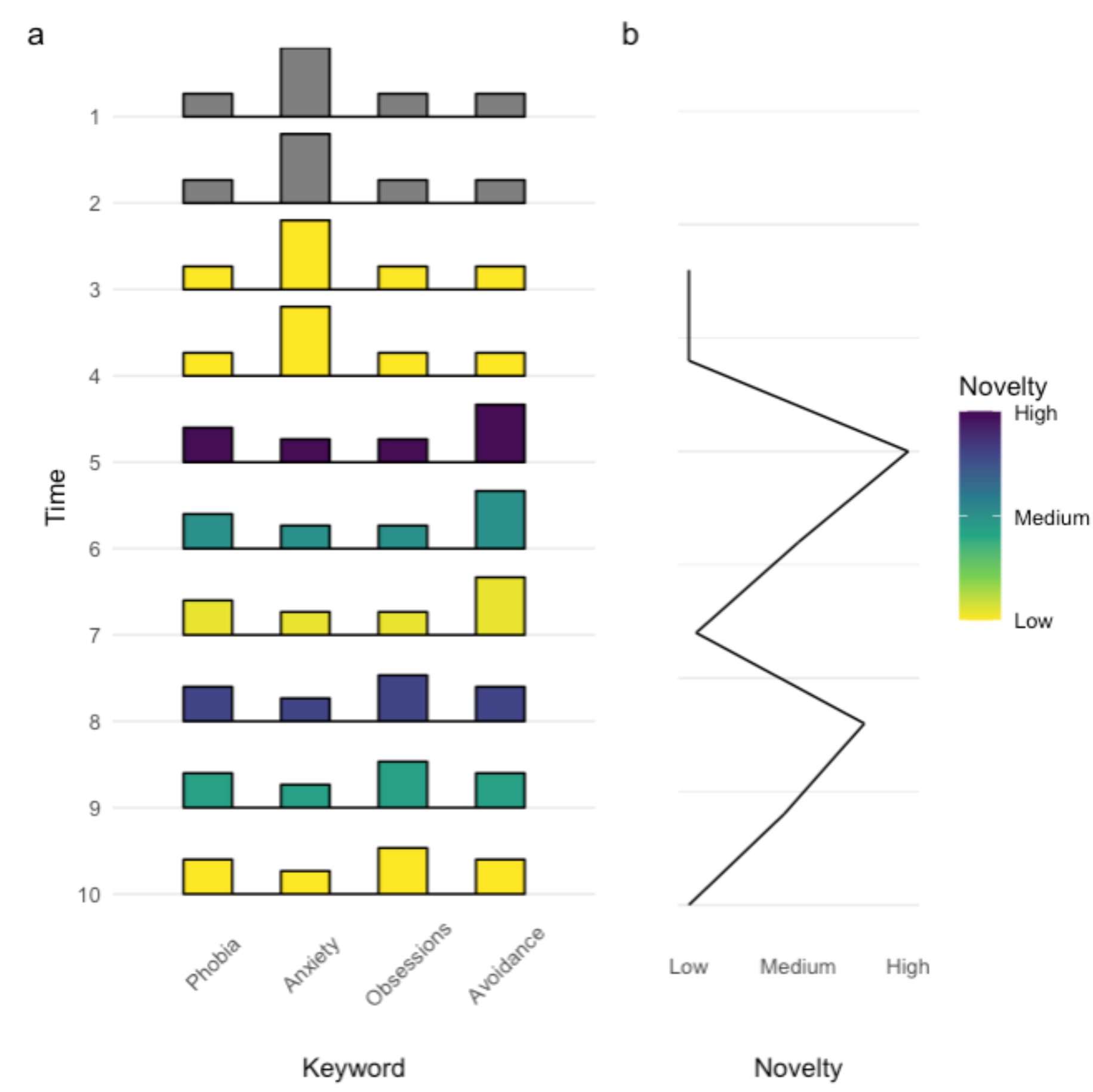
Example of novelty calculation with a window size of 2. (a) shows the distribution shift over time of four keywords from the F4 category. The distribution of the four keywords at each timepoint is represented with a histogram, with colour indicating novelty levels. (b) shows novelty at the different time points. Major shifts such as from time 4 to 5 lead to high novelty, which gradually decreases as the distribution becomes stable. The larger the shift in distribution, the greater the novelty. Novelty cannot be calculated for the first two points as we have defined the window size to be two.

#### Mean note length

We calculated the mean note length as the mean number of tokens in each document. A token is a meaningful segment of text and includes whitespace-separated words, punctuation and more^35^. For the present study, the documents were tokenized using spaCy’s Danish tokenization module^36^.

#### Automated Readability Index

The Automated Readability Index (ARI) is a measure of the readability of a text, designed to estimate the grade level required to comprehend the text^32^. The equation below shows the formula for calculating the ARI.

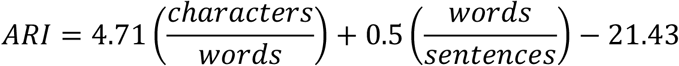

#### Dependency Distance

We define dependency distance as the number of words between a word and its headword. The mean dependency distance is the average dependency distance of a document or sentence, as visualized in Figure 6. The dependency tree is obtained by performing a dependency parse using spaCy, and dependency distance was calculated using *textdescriptives*^29^.

**Figure 6:**
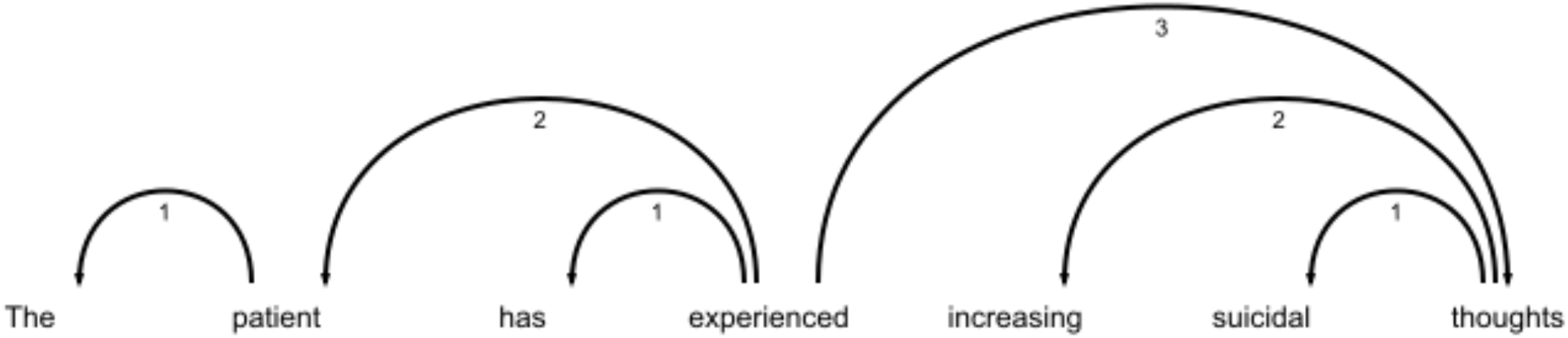
Calculation of dependency distance. Numbers on arrows indicate the distance from the head to the word. Note that ‘experienced’ has no arrows pointing to it, as it is the root of the sentence. The dependency distance of ‘experienced’ is therefore zero. The mean dependency distance of the sentence is (1+2+1+0+2+1+3) / 7 = 1.43.

#### Changepoint detection

All time series (83) were analysed for multiple change points in both mean and variance using the Pruned Exact Linear Time (PELT) algorithm^37^. Specifically, the time series were 1) one for each of the 17 note types for each for the four descriptive statistics (mean number of tokens, mean dependency distance, ARI, and proportion of notes (68 in total), 2) three time series for the aggregated notes (mean number of tokens, mean dependency distance, and ARI), and 3) a time series for the novelty of each of the 10 ICD-10 diagnostic categories of words describing psychopathology, one for the novelty of words used in the mental state examination, and one for the novelty of all words (aggregate) describing psychopathology (12 in total). PELT automatically finds the appropriate number of change points by iteratively searching for new changepoints at each data split. This makes PELT particularly suitable for tasks without prior information on the location and number of changepoints. Variables were detrended by differencing with a lag of one time step and z-scored before input to PELT using the default parameters of the *cpt*.*meanvar* function (penalty=“MBIC”, minseqlen=2). Changepoint detection was conducted in R v3.6.1^38^ using the *tidyverse v1*.*3*.*1*^39^ and *changepoint v2*.*2*.*2*^40^ packages. Given the 83 time series with a datapoint for each quarter for nine years (2013 Q1 - 2021 Q4), the number of data points (and hence the number of possible changepoints) was 4 × 9 × 83 = 2988.

#### Stability of the use of words describing psychopathology

The analyses of the stability in the use of the 365 words describing psychopathology (Figure 2) were conducted by identifying the words that had the largest relative change from the mean of the two preceding time points to the time point of interest. The relative change in word usage was quantified as the ratio between the current novelty value and the mean of the two previous points. The divergence metric in the novelty equation is sensitive to relative changes in proportions as opposed to absolute changes, i.e., a change in proportional usage of a keyword from 0.4 to 0.2 will have the same impact on novelty as a change from 0.1 to 0.05. Hence, words that are used rarely are more likely to have large fluctuations in relative proportions and are therefore stronger drivers of novelty than more commonly used words.

### Ethics

This study was carried out to ensure the validity/stability of the data used for studies based on the PSYCOP cohort^18^. The use of electronic health records from the Central Denmark Region was approved by the Central Denmark Region Legal Office per the Danish Health Care Act §46, Section 2. According to the Danish Committee Act, ethical review board approval is not required for studies based solely on data from electronic health records (waiver for this project: 1-10-72-1-22). All data were processed and stored in accordance with the European Union General Data Protection Regulation, and the project is registered on the internal list of research projects having the Central Denmark Region as the data steward.

## Data Availability

The data used for the study is not publicly available as it contains personally sensitive information. All code can be found online at https://github.com/Aarhus-Psychiatry-Research/lexical-stability

## DATA AVAILABILITY

In accordance with Danish law for the protection of privacy, the data used for this study is only available for research projects conducted by employees in the Central Denmark Region upon approval from the Legal Office under the Central Denmark Region (in accordance with the Danish Health Care Act §46, Section 2).

## CODE AVAILABILITY

All code used for extracting and analysing the data can be found on the following Github repository: https://github.com/Aarhus-Psychiatry-Research/lexical-stability. While not all steps can be reproduced due to data privacy, we have provided the aggregated data files used for the changepoint analyses.

## ACKNOWLEDGEMENTS

The authors thank Bettina Nørremark from Aarhus University Hospital – Psychiatry for her assistance with data extraction.

## AUTHOR CONTRIBUTIONS

Conception and design: all authors. Funding obtainment: S.D.Ø. Provision of study data: S.D.Ø. and A.A.D. Data analysis: L.H. K.C.E. Interpretation: All authors. Manuscript writing: L.H., E.P, M.B. and S.D.Ø. Revision of manuscript for important intellectual content: K.C.E., A.A.D., K.L.N. Final approval of the manuscript: all authors.

## ETHICAL APPROVAL

This study was carried out to ensure the validity/stability of the clinical notes used for studies based on the PSYCOP cohort. The use of electronic health records from the Central Denmark Region was approved by the Central Denmark Region Legal Office per the Danish Health Care Act §46, Section 2. According to the Danish Committee Act, ethical review board approval is not required for studies based solely on data from electronic health records (waiver for this project: 1-10-72-1-22). The data were stored and processed in accordance with the European Union General Data Protection Regulation. The project is registered on the internal list of research projects having the Central Denmark Region as the data steward.

