## Supplementary materials for "Lexical Stability of Psychiatric Clinical Notes from Electronic Health Records over a Decade"

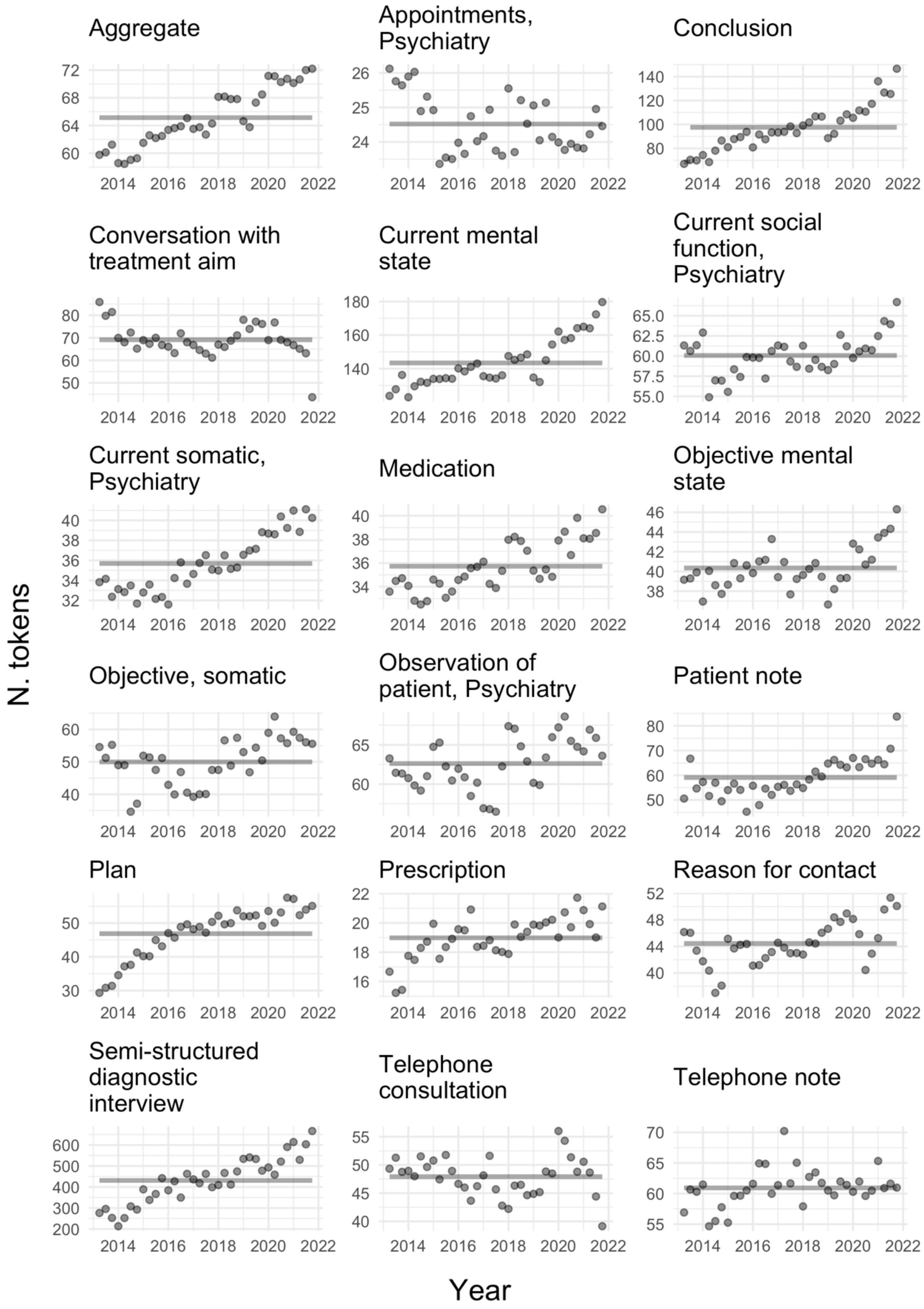


**Supplementary Figure 1:** Mean number of tokens per note type with overlaid changepoint detection.

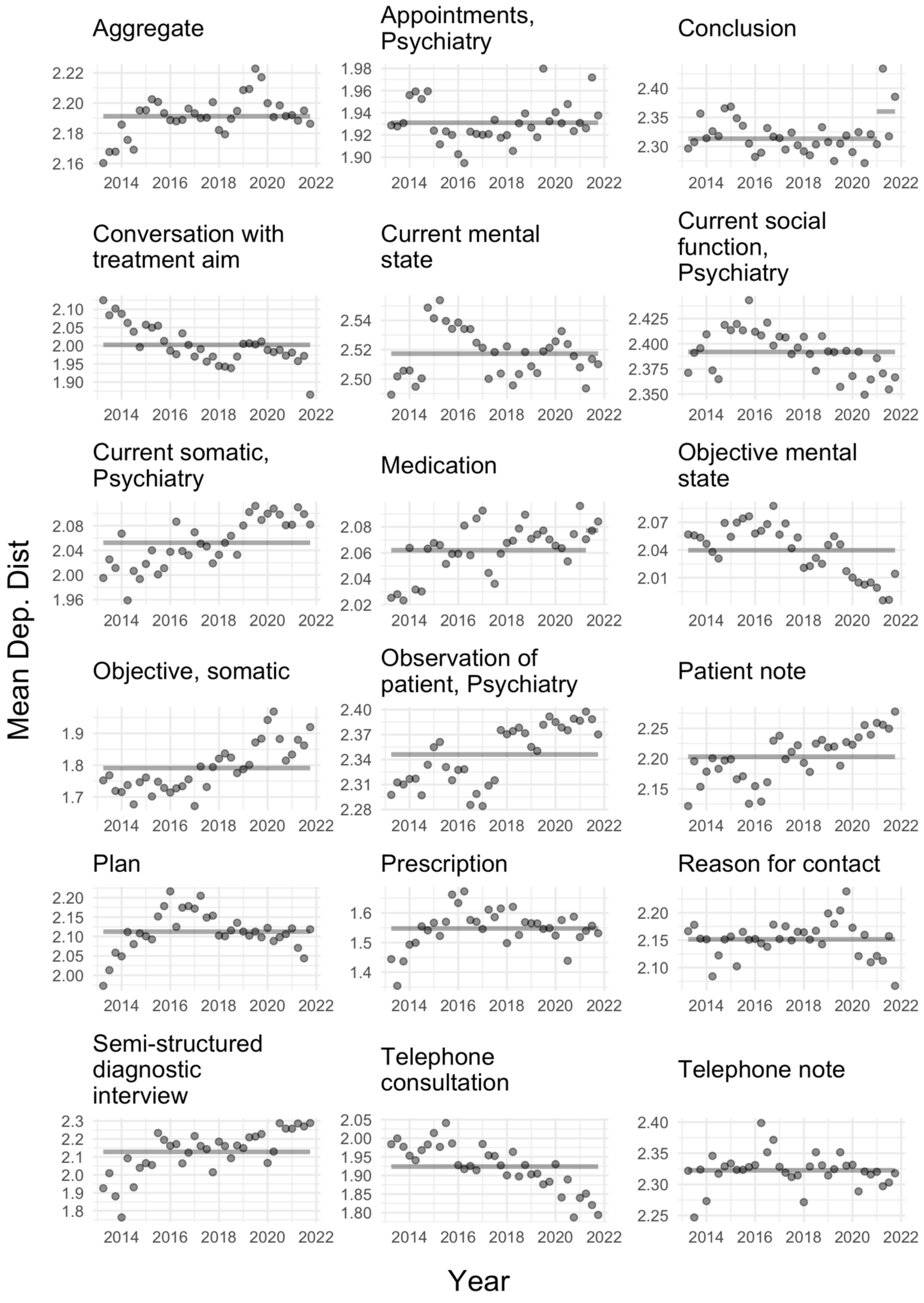


**Supplementary Figure 2:** Mean dependency distance per note type with overlaid changepoint detection.

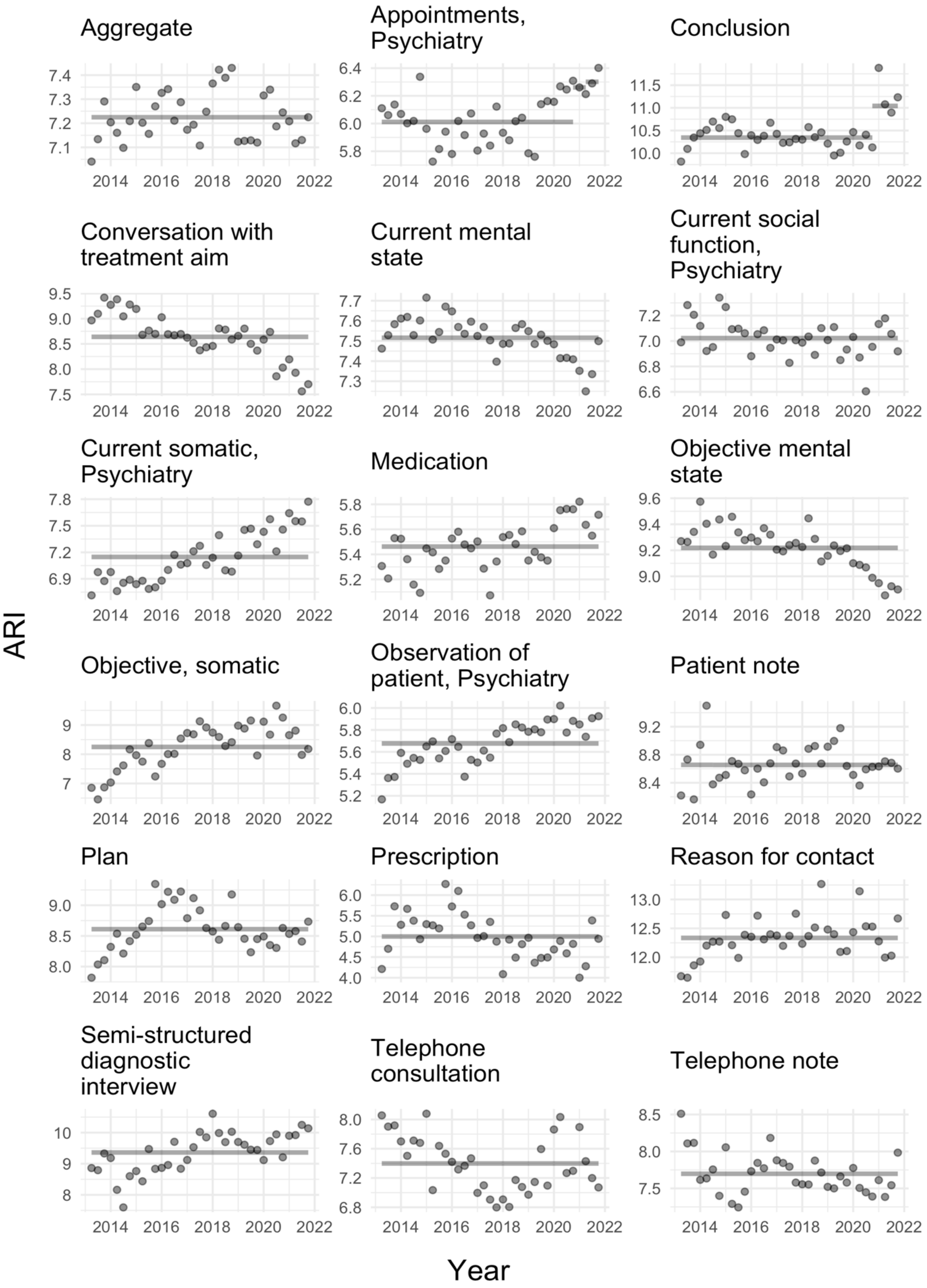


**Supplementary Figure 3:** Mean Automated Readability Index per note type with overlaid changepoint detection.

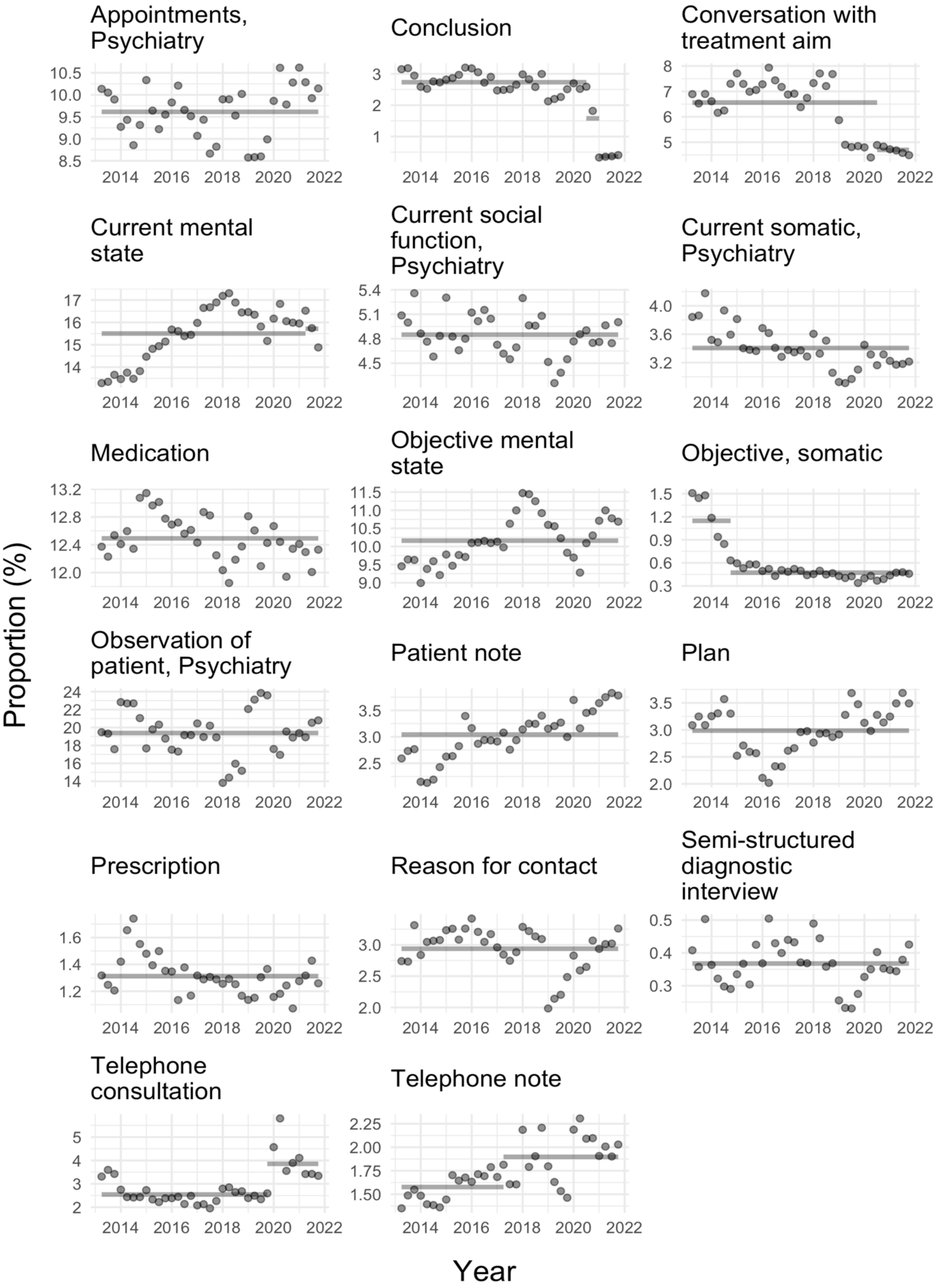


**Supplementary Figure 4:** Mean proportion of all notes per note type with overlaid changepoint detection. Note, proportion of notes for "Aggregate" cannot be calculated and is therefore not shown.

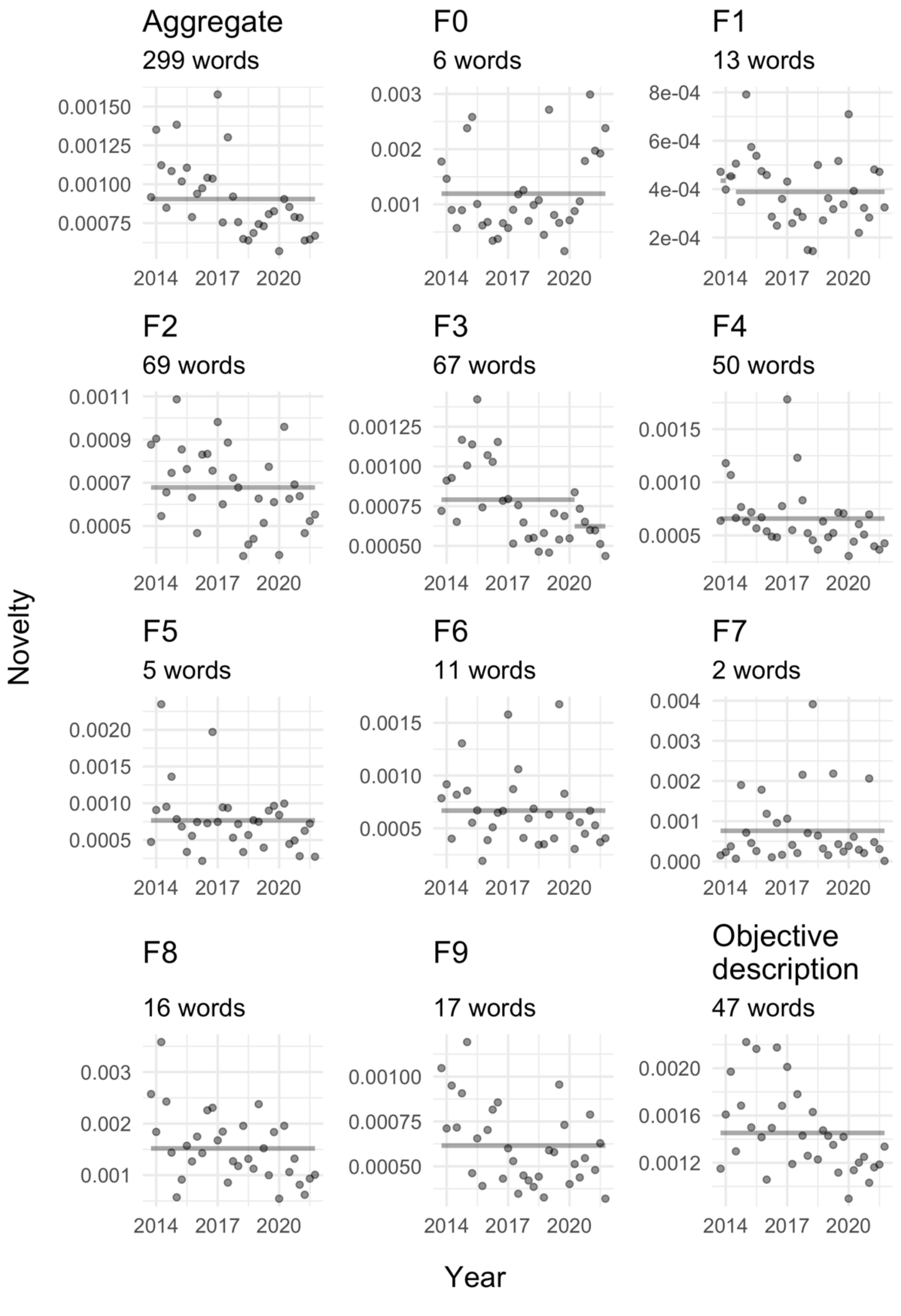


**Supplementary Figure 5:** Novelty over time for each category of words describing psychopathology. Grey lines indicate changepoints with lines showing the mean of the segment. The number of words in each category is shown in the subtitle of each plot.

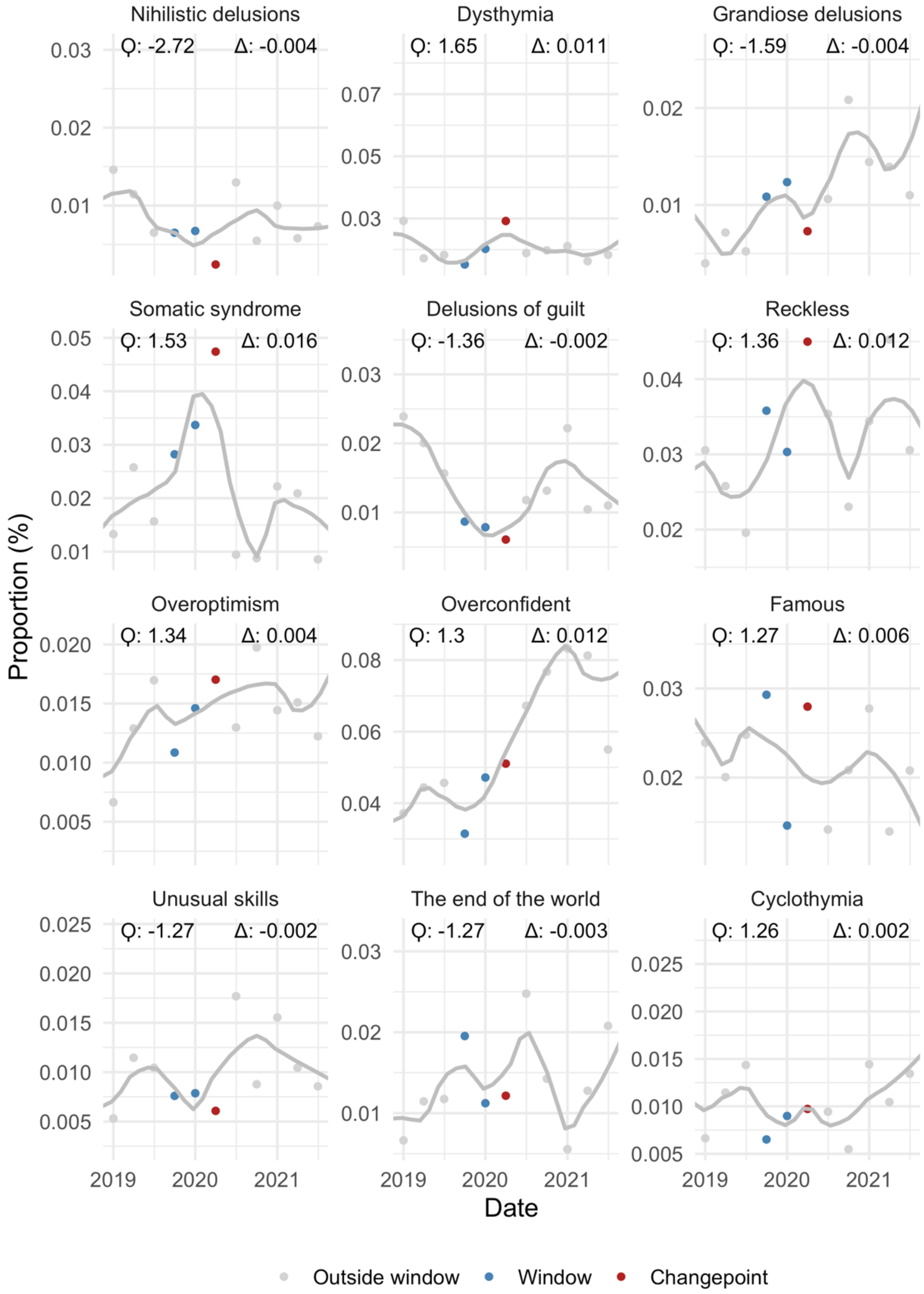


**Supplementary Figure 6**: The 12 F3 – Mood disorders related words with the largest relative change in proportion from 2019 Q4 + 2020 Q1 to 2020 Q2. Ϙ shows the quotient difference between the mean of the two previous points (window) to the point of interest, i.e., 2020 Q2, and Δ shows the absolute difference between the mean of the two previous points to the point of interest. The y-axis indicates how large a percentage of all words the keyword in question makes up.

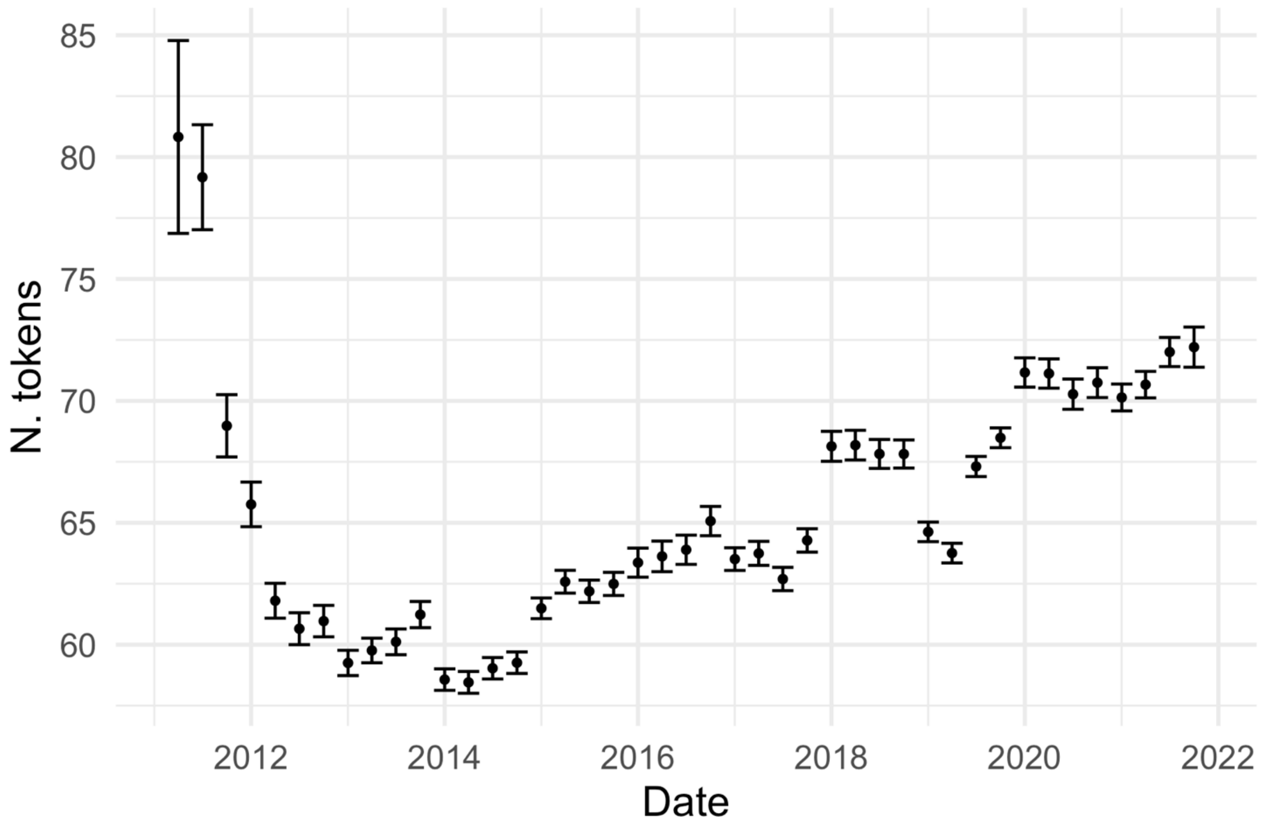


**Supplementary Figure 7: Mean number of tokens per note on the aggregate level including 2011 and 2012.** Error bars indicate +- the standard error of the mean. Data before approximately Q2 2012 have higher variance and significantly higher means.

**Supplementary Table 1: Description of the content of each of the note types.**

| Danish name | English name | Description |
|---|---|---|
| Aftaler, Psykiatri | Appointments, Psychiatry | Description of concrete care- and treatment-related appointments and agreements with and about the patient. |
| Aktuelt socialt, Psykiatri | Current social, Psychiatry | Description of the patient's current social situation: relationship to the family, civil status, residential-, occupational-, and economic conditions, and contact with the social services. Documentation of ongoing treatment plans in relation to the patient's social relationships. |
| Aktuelt psykisk | Current mental state | Description of the development, progress, and current status of the patient's mental illness. |
| Aktuelt somatisk, Psykiatri | Current somatic, Psychiatry | Description of the patient's current and chronic somatic illnesses and symptoms. Information on current treatment in relation to the patient's physical condition. |
| Journalnotat | Journal note | Treatment notes that cannot be meaningfully described in another note type. |
| Konklusion | Conclusion | Aggregation and interpretation of all findings and decisions based on an interview (e.g in relation to an outpatient visit or during an inpatient stay). |
| Kontaktårsag | Reason of contact | The reason for the patient's in- or outpatient treatment course. |
| Medicin | Medication | Documentation of indications and reasons for prescription of medication, adjustments in dose of |

| | | |
|---|---|---|
| | | medication, actions in relation to administration of medication. |
| Objektivt psykisk | Objective mental state | Objective assessment of the patient's mental state, including state of consciousness, orientation, intelligence, psychomotor function, mood, delusions, psychotic symptoms, etc. |
| Objektivt, somatisk | Objective, somatic | Documentation of somatic assessments/examinations. |
| Observation af patient, Psykiati | Observation of patient, Psychiatry | Description of an inpatient's mental symptoms, behaviour, reactions towards relatives, other patients, staff, etc. |
| Ordination | Prescription of examinations | Prescription of examinations such as blood pressure measurement, pulse measurement, weighing, medical imaging, laboratory tests, etc. |
| Plan | Plan | Acute or short-term plans for the patient. |
| Samtale med behandlingssigte | Conversation with treatment aim | Documentation of the conversation's purpose and attendees. |
| Semistruktureret diagnostisk interview | Semi-structured diagnostic interview | Registration that a semi-structured diagnostic interview has been conducted. Description of the method and documentation of result and conclusion. |
| Telefonkonsultation | Telephone consultation | Documentation of telephone communication with the patient or the patient's guardian as part of treatment or monitoring of disease progress. |
| Telefonnotat | Telephone note | Documentation of telephone conversations with a patient or the |

| | | |
|---|---|---|
| | | patient's guardian of non-clinical character. |

**Supplementary Table 2: Psychopathological keywords used for the analysis.** The table shows the keywords used for the novelty analysis in both Danish and English. An X in the 'excluded' column indicates that the keyword was not included in the analysis due to too few occurences. An "/" in the name indicates that the counts for the two words have been added.

| Diagnostic chapter | Danish | English | Excluded |
|---|---|---|---|
| **F0** | - | - | |
| | Demens | Dementia | |
| | Vaskulær demens | Vascular dementia | |
| | Alzheimer | Alzheimer | |
| | Lewy body | Lewy body | |
| | Delir | Delirium | |
| | Delirøs | Delirous | |
| **F1** | - | - | |
| | Misbrug | Drug abuse | |
| | Trang | Urge | |
| | Craving | Craving | |
| | Svigtende kontrol | Loss of control | X |
| | Abstinenssymptomer | Withdrawal symptoms | |
| | Tolerans | Tolerance | |
| | Risikofyldt adfærd | Risky behaviour | X |
| | Alkohol | Alcohol | |
| | Opioder | Opiods | X |
| | Cannabis | Cannabis | |
| | Hash | Hash | |
| | Cannabinoider | Cannabinoids | |
| | Sedativa | Sedatives | |
| | Hypnotika | Hypnotics | |
| | Kokain | Cocaine | |
| | Centralstimulerende | Central stimulant | |
| | Hallucinogener | Hallucinogens | X |
| **F2** | - | - | |
| | Skizofreni | Schizophrenia | |
| | Hebefren | Hebephrenic | |
| | Skizotypi | Schizotypal | |
| | Paranoid | Paranoid | |
| | Paranoia | Paranoia | |
| | Psykose | Psychosis | |
| | Psykotisk | Psychotic | |
| | Skizoaffektiv / skizo-affektiv | Skizoaffective | |
| | Positive symptomer | Positive symptoms | |
| | Auditive hallucinationer | Auditory hallucinations | |

| | | | |
|---|---|---|---|
| | Hørehallucinationer | Hearing hallucinations | |
| | Verbale | Verbal | |
| | Non-verbale | Non-verbal | |
| | 2. person / andenpersons | 2nd person / second person | |
| | 3. person / tredjepersons | 3rd person / third person | |
| | Kommenterende | Commenting | |
| | Diskuterende | Discussing | |
| | Depressiv | Depressive | |
| | Expansive | Expansive | X |
| | Dissociative | Dissociative | |
| | Hypnagoge | Hypnogogic | |
| | Hypnopompe | Hypnopompic | |
| | Stemmer | Voices | |
| | Devaluerende | Devaluating | |
| | Synshallucinationer | Visual hallucinations | |
| | Uformede | Unformed | |
| | Formede | Formed | |
| | Sceniske | Scenic | |
| | Interne | Internal | |
| | Eksterne | External | |
| | Lugthallucinationer | Olfactory hallucinations | |
| | Taktile hallucinationer | Tactile hallucinations | |
| | Somatiske hallucinationer | Somatic hallucinations | |
| | Berøringshallucinationer | Hallucinations of touch sensations | |
| | Seksuelle hallucinationer | Sexual hallucinations | |
| | Legemlige fornemmelser | Bodily sensations | X |
| | Tankeforstyrrelser | Thought disorders | |
| | Tankefradrag | Thought withdrawal | |
| | Tankepåføring | Thought insertion | |
| | Tankeudspredning | Thought broadcasting | |
| | Tankehørlighed | Loud thoughts | |
| | Tanke-ekko | Thought echo | |
| | Styringsoplevelser | Delusions of control | |
| | Påførte handlinger | Replaced control of actions | |
| | Fremmed vilje | Alien will | X |
| | Påført tale | Replaced control of voice | |
| | Påført skriven | Replaced control of handwriting | X |

| | | | |
|---|---|---|---|
| | Påførte følelser | Replaced control of affect | |
| | Fremmed magt | Alien force | X |
| | Påførte impulser | Replaced control of impulses | X |
| | Vrangforestillinger | Delusions | |
| | Vrangstemning | Delusional mood | |
| | Selvhenførende vrangforestillinger | Delusions of reference | |
| | Mistydning | Misinterpretation | |
| | Primær vrangsansning | Delusional perception | |
| | Persekutoriske vrangforestillinger | Delusions of persecution | |
| | Forfølgelsesforestillinger | Delusions of persecution | |
| | Forfulgt | Persecuted | |
| | Forgifte | Poison | |
| | Skade dig | Hurt you | X |
| | Sammensværgelsesforestillinger | Delusions of conspiracy | X |
| | Komplot | Plot | |
| | Religiøse forestillinger | Religious delusions | |
| | Guddommelig | divine | |
| | Religiøse budskaber | Religious messages | X |
| | Overnaturlige forklaringer | Supernatural explanations | X |
| | Okkult | Occult | |
| | Telepati | Telepathy | |
| | Seksuelle forestillinger | Sexual delusions | X |
| | Jalousiforestillinger | Delusional jealousy | X |
| | Fantastiske forestillinger | Fantasy delusions | X |
| | Bizarre vrangforestillinger | Bizarre delusions | |
| | Førsterangssymptomer | First rank symptoms | |
| | Katatoni / kataton | Catatonia / catatonic | |
| | Negative symptomer | Negative symptoms | |
| | Koncentrationsbesvær | Concentration problems | |
| | Tab af følelser | Loss of emotions | |
| | Social tilbagetrækning | Social withdrawal | |
| | Formålsløs adfærd | Purposeless behaviour | X |
| | Passivitet | Passitivity | X |
| | Manglende fremdrift | Lack of progress | |
| | Derealisation | Derealisation | |
| | Depersonalisation | Depersonalisation | |
| | Perceptionsforstyrrelse | Perception disorder | |

| | | | |
|---|---|---|---|
| | Fremmedfølelse | Alienation | |
| **F3** | - | - | |
| | Bipolar affektiv sindslidelse | Bipolar affective disorder | |
| | Bipolar | Bipolar | |
| | Mani / manisk | Mania / manic | |
| | Hypomani / hypomanisk | Hypomania / hypomanic | |
| | Expansivt stemningsleje | Expansive mood | X |
| | Eretisme | Eretism | X |
| | Øget energi | Increased energy | |
| | Hyperaktiv | Hyperactive | |
| | Tankepres | Pressing thoughts | |
| | Tankeflugt | Racing thoughts | |
| | Talepres | Pressure of speech | |
| | Taletrang | Overtalkativeness | |
| | Grandios | Grandiose | |
| | Grandiose vrangforestillinger | Grandiose delusions | |
| | Megalomane | Megalomanic | |
| | Særlig mission | Special mission | X |
| | Usædvanlige krafter | Unusual power | X |
| | Usædvanlige evner | Unusual skills | |
| | Berømt | Famous | |
| | Letafledlighed | Ease of diversion | X |
| | Købetrang | Urge for spending | |
| | Over-optimisme | Overoptimism | |
| | Kompromitterende adfærd | Socially embarassing behavior | |
| | Hæmningsløs | Unhibited | |
| | Overmodig | Overconfident | |
| | Overfamiliaritet | Over-familiarity | |
| | Ukritisk adfærd | Uncritical behaviour | X |
| | Hensynsløs | Reckless | |
| | Uansvarlig adfærd | Irresponsible behavor | |
| | Nye interesser | Novel interests | |
| | Distraktibilitet | Distractibility | X |
| | Nedsat søvnbehov | Decreased need for sleep | |
| | Øget seksualtrang | Increased sexual urge | |
| | Øget sex-drift | Increased sexual drive | X |
| | Depression | Depression | |
| | Nedtrykthed | Sadness | |
| | Nedsat stemningsleje | Depressed mood | |
| | Nedsat humør | Depressed mood | |

| | | | |
|---|---|---|---|
| | Nedsat energi | Loss of energy | |
| | Trætbarhed | Fatigue | |
| | Nedsat interesse | Loss of interests | |
| | Nedsat lyst | Decreased desire | |
| | Interessetab | Loss of interests | |
| | Håbløshed | Hopelessness | |
| | Hæmning | Inhibition | |
| | Rumination | Rumination | |
| | Pessimisme | Pessimism | |
| | Grådtendens | Tearfullnees | |
| | Dødstanker | Thoughts of death | |
| | Selvmordstanker | Suicidal ideation | |
| | Selvmord | Suicide | |
| | Selvmordforsøg | Suicide attempt | |
| | Selvskade | Self-harm | |
| | Nedsat selvtillid | Loss of self-confidence | |
| | Selvbebrejdelser | Self-Blame | |
| | Vægttab | Loss of weight | |
| | Nedsat libido | Loss of libido | |
| | Nedsat sexlyst | Loss of libido | |
| | Nedsat appetit | Lack of appetite | |
| | Initiativløshed | Lack of iniative | |
| | Søvnløs | Insomnia | |
| | Søvnbesvær | Insomnia | |
| | Tidlig opvågen | Early waking | |
| | Tidlig opvågning | Early waking | |
| | Melankoliform | Somatic syndrome | |
| | Depressive vrangforestillinger | Delusions in the context of depression | |
| | Skyldsforestillinger | Delusions of guilt | |
| | Skyldfølelse | Feeling of guilt | |
| | Selvforringende | Self-deteriorating | |
| | Dårligt menneske | Bad person | |
| | Uduelig | Incompetent | |
| | Betydningsløs | Insignificant | X |
| | Forarmningsforestillinger | Delusions of impoverishment | X |
| | Ruineret | Ruined | X |
| | Verdens undergang | The end of the world | |
| | Hypokondre forestillinger | Hypochondrical delusions | X |
| | Nihilistiske forestillinger | Nihilistic delusions | |
| | Rådne | Rot | |
| | Cotard's syndrom | Cotard syndrome | X |

| | | | |
|---|---|---|---|
| | Dysmorfofobe forestillinger | Delusions of dysmorphophobia | X |
| | Derealisationsvrangforestillinger | Delusions of derealisation | |
| | Capgra's syndrom | Capgra's syndrome | X |
| | Cyklothymi | Cyclothymia | |
| | Dysthymi | Dysthymia | |
| **F4** | - | - | |
| | Fobi | Phobia | |
| | Agorafobi | Agoraphobia | |
| | Socialfobi | Social phobia | |
| | Social angst | Social Anxiety | |
| | Enkelfobi | Specific phobia | |
| | Generaliseret angst | Generalized anxiety | |
| | Belastningsreaktion | Acute stress reaction | |
| | Tilpasningsreaktion | Adjustment disorders | |
| | Dissociativ | Dissociative | |
| | Somatoform | Somatoform | |
| | Bekymringstendens | Worry | |
| | Anspændt | Tense | |
| | Generel muskelspænding | General muscle tension | X |
| | Angst | Anxiety | |
| | Bange | Afraid | |
| | Undgåelse | Avoidance | |
| | Panikanfald | Panic attack | |
| | Panikangst | Panic disorder | |
| | Hjertebanken | Palpitations | |
| | Smerter i brystet | Chest pain | |
| | Trykken for brystet | Chest discomformt | |
| | Åndedrætsbesvær | Difficulty breathing | |
| | Kvælningsfornemmelse | Feeling of choking | |
| | Kvalme | Nausea | |
| | Maveuro | Abdominal distress | |
| | Hedeture | Hot flushes | |
| | Kuldegysninger | Cold chills | |
| | Sveden | Sweating | |
| | Mundtørhed | Dry mouth | |
| | Rysten | Trembling | |
| | Sitren | Tingling sensations | |
| | Dødsangst | Fear of dying | |
| | Synkebesvær | Difficulty with swallowing | |

| | | | |
|---|---|---|---|
| | Klump i halsen | Sensation of a lump in the throat | |
| | Uvirkelighedsfølelse | | |
| | Undgåelsesadfærd | Avoidance behavior | |
| | Paræstesier | Paraesthesias | |
| | Føleforstyrrelser | Sensory disturbances | |
| | PTSD | PTSD | |
| | Posttraumatisk belastningsreaktion | Post-traumatic stress disorder | |
| | Flashbacks | Flashbacks | |
| | Mareridt | Nightmare | |
| | Hypervigilitet | Hypervigilant | |
| | Katastrofe | Catastrophe | |
| | Traume | Trauma | |
| | Obsessiv-kompulsiv tilstand | Obsessive-Compulsive Disorder | |
| | OCD | OCD | |
| | Tvangstanker | Obsessions | |
| | Tvangshandlinger | Compulsions | |
| | Fugue | Fugue | |
| | Tranceoplevelser | Experiences of trance | X |
| | Besættelsesoplevelse | Possession disorders | |
| **F5** | - | - | |
| | Spiseforstyrrelse | Eating disorders | |
| | Anorexi | Anorexi | |
| | Anoreksi | Anorexi | |
| | Anorexia nervosa | Anorexia nervosa | |
| | Bulimi | Bulimia | |
| **F6** | - | - | |
| | Personlighedsforstyrrelse | Personality disorders | |
| | Paranoid | Paranoid | |
| | Skizoid | Schizoid | |
| | Dyssocial | Dissocial | |
| | Emotionel ustabil personlighedsstruktur | Emotionally unstable personality disorder | |
| | Borderline | Borderline | |
| | Histrionisk | Histrionic | |
| | Tvangspræget | Anankastic | |
| | Ængstelig | Anxious | |
| | Evasiv | Avoidant | |
| | Dependent | Dependent | |
| **F7** | - | - | |
| | Mental retardering | Mental retardation | |
| | Lav IQ | Low IQ | |

| | | | |
|---|---|---|---|
| **F8** | - | - | |
| | Gennemgribende udviklingsforstyrrelser | Pervasive developmental disorders | |
| | Infantil autisme | Childhood autism | |
| | Autisme | Autism | |
| | Asperger | Asperger | |
| | Manglende blikkontakt | Lack of Eye-to-eye gaze | |
| | Nedsat blikkontakt | Decreased eye-to-eye gaze | |
| | Nedsat mimik | Decreased facial expression | |
| | Nedsat gestik | Decreased gesture | |
| | Stereotyp adfærd | Stereotyped behavior | |
| | Repetitivt | Repetitive | |
| | Impressivt sprog | Receptive language | X |
| | Expressivt sprog | Expressive language | X |
| | Nedsat spontan tale | Lack of spontaneous speech | X |
| | Idiosynkratisk sprogbrug | Idiosyncratic use of words or phraeses | |
| | Indsnævrede interesser | Restricted patterns of interests | |
| | Afvigende socialt samspil | Abnormal social interaction | |
| | Afvigende kommunikation | Abnormal communication | |
| | Nedsat empati | Lack of empathy | |
| | Nedsat situationsfornemmelse | Lack of modulation of behavior in a social context | |
| | Emotionel dysregulering | Emotional dysregulation | X |
| **F9** | - | - | |
| | Opmærksomhedsforstyrrelser | Attention disorders | |
| | Hyperkinetiske forstyrrelser | Hyperkinetic disorders | |
| | Hyperkinetisk | Hyperkinetic | |
| | ADHD | ADHD | |
| | ADD | ADD | |
| | Koncentrationsbesvær | Concentration difficulties | |
| | Skødesløse fejl | Careless mistakes | |
| | Besvær med at planlægge | Difficulties in planning | X |

| | | | |
|---|---|---|---|
| | Mister ting | Looses items | |
| | Motorisk aktivitet | Motor activity | |
| | Hyperaktiv | Hyperactive | |
| | Impulsiv | Impulsive | |
| | Impulsivitet | Impulsivity | |
| | Afbryder | Interrupts | |
| | Søvnbesvær | Sleeplessness | |
| | Adfærdsforstyrrelse | Conduct disorder | |
| | Tics | Tics | |
| | Tourettes | Tourettes | |
| **Mental status examination** | - | - | |
| | Perseveration | Perseveration | |
| | Konfabulation | Confabulation | X |
| | Amnesi | Amnesia | |
| | Opmærksomhedssvækkelse | Lack of concentration | X |
| | Desorientering | Desorientated | |
| | Apati | Apathy | |
| | Afasi | Aphasia | |
| | Træghed | Inertia | |
| | Opfattelsesbesvær | Perception difficulties | X |
| | Apraksi | Apraxia | |
| | Stupor | Stupor | |
| | Initiativløshed | Lack of iniative | |
| | Monoton stemme | Monotone voice | |
| | Manglende øjenkontakt | Lack of eye-to-eye gaze | |
| | Letafledelig | Distractible | |
| | Rastløs | Restless | |
| | Agiteret | Agitated | |
| | Hyperaktiv | Hyperactive | |
| | Påfaldende udseende eller påklædning | Striking appearance or clothing | X |
| | Usammenhængende adfærd | Incoherent behavior | X |
| | Excentrisk adfærd | Excentric behavior | X |
| | Selvforsømmelse | Self-neglect | |
| | Dårlig hygiejne | Bad hygiene | |
| | Kataton | Catatonic | |
| | Negativisme | Negativism | X |
| | Automatisk lydighed | Automatic obedience | X |
| | Katalepsi | Catalepsy | X |
| | Flexibilitas cerea | Flexibilitas cera | X |
| | Abnorm legemsholdning | Abnormal posture | X |
| | Stirren | Stare | |

| | | | |
|---|---|---|---|
| | Grimasseren | Grimaces | |
| | Bevægelsesstereotypier | Motor stereotypes | |
| | Stereotypier | Stereotypes | |
| | Løftet stemningsleje | Elevated mood | |
| | Euforisk | Euphoric | |
| | Irritabel | Irritable | |
| | Forvirring / forvirret | Confusion / confused | |
| | Emotionel labilitet | Emotional lability | |
| | Mistroisk | Suspiscious | |
| | Histrionisk affekt | Histrionic affect | X |
| | Ambivalens / ambivalent | Ambivalence / ambivalent | |
| | Indsnævret affekt | Restricted affect | X |
| | Inadækvat affekt | Inadequate affect | |
| | Inkongruent affekt | Incongruity of affect | |
| | Aggressiv | Aggressive | |
| | Affektaffladning | Flattening of affect | |
| | Affektlabil | Lability of mood | |
| | Manglende emotionelt respons | Lack of emotional respons | X |
| | Latenstid | Latency | |
| | Omstændelig | Elaborate | |
| | Springende tankegang | Flight of ideas | |
| | Associerer frit | Associating freely | X |
| | Frit associerende | Freely associating | |
| | Tankeflugt | Racing thoughts | |
| | Usammenhængende tankegang | Incoherent thoughts | |
| | Usamlet | Incoherent | |
| | Kaotisk tankegang | Chaotic thoughts | |
| | Inkoherent tale | Incoherent speech | X |
| | Neologismer | Neologisms | |
| | Metaforisk tale | Metaforical speech | X |
| | Metonymier | Metonymi | X |
| | Vaghed | Poverty of content of speech | |
| | Mutisme | Mutism | |
| | Mumler | Mumbling | |
| | Perseverationstendens | Perseveration | |
